# Short Term Reduction in the Odds of Testing Positive for SARS-CoV-2; a Comparison Between Two Doses and Three doses of the BNT162b2 Vaccine

**DOI:** 10.1101/2021.08.29.21262792

**Authors:** Tal Patalon, Sivan Gazit, Virginia E. Pitzer, Ottavia Prunas, Joshua L. Warren, Daniel M. Weinberger

## Abstract

With the evidence of waning immunity of the BNT162b2 vaccine, a national third dose vaccination campaign was initiated in Israel during August 2021; other countries have announced their intention to administer a booster shot as well. Leveraging data from Maccabi Healthcare Services, we conducted a preliminary retrospective study aimed at evaluating initial short-term effectiveness of a three dose versus a two dose regimen against infection due to the Delta variant of SARS-CoV-2, using two complementary approaches; a test-negative design and a matched case-control design. We found that 7-13 days after the booster shot there is a 48-68% reduction in the odds of testing positive for SARS-CoV-2 infection and that 14-20 days after the booster the marginal effectiveness increases to 70-84%. Further studies are needed to determine the duration of protection conferred by the third dose and its effect on severe disease.

## Introduction

The short-term effectiveness of a two-dose regimen of the BioNTech/Pfizer mRNA BNT162b2 vaccine against severe acute respiratory syndrome coronavirus 2 (SARS-CoV-2) was demonstrated both in clinical trials^1,2^ and real-world settings.^3,4^ However, long-term effectiveness remains undetermined, as evidence of waning immunity has surfaced both in terms of antibody dynamics over time^5–7^ and protection against infection.^8^

The Delta (B.1.617.2) variant of SARS-CoV-2 was the dominant strain in Israel during the summer of 2021 and caused a rapidly growing number of cases, many in vaccinated individuals.^9^ A national third-dose (booster) vaccination campaign was initiated in Israel, and as of August 24, 2021, a booster dose was recommended for those aged 30 and over as well as for high-risk populations.^10^ By August 27, 2021, over 1.9 million individuals had received a booster shot.^9^ Current guidelines state that a minimum interval of 5 months since receipt of the second dose is required to be eligible for the booster dose. According to recent announcements, booster shots will be offered to individuals in the United States and United Kingdom in the coming months.

Few studies on a third-dose regimen have been published to date, all of which are in high-risk populations, such as transplant patients.^11,12^ These reported adequate antibody responses, though follow-up time was short and cohort sizes small. Recently, Pfizer and BioNTech released a statement regarding a reported on-going trial, in which a booster was administered 6 months after the second dose. The report indicated 5 to 10 times higher neutralization titers after the third dose, though neither data nor manuscript have yet to be published.^13^

To this end, we conducted a preliminary retrospective study aimed at evaluating initial short-term marginal effectiveness of the third dose of the BNT162b2 vaccine compared to a two-dose regimen, leveraging data from Maccabi Healthcare Services (MHS), Israel’s second largest Health Maintenance Organization, which covers 2.5 million members.

We used two complementary approaches to evaluate to marginal benefit of the third dose, both previously undertaken to evaluate SARS-CoV-2 vaccine effectiveness, each with its own limitations: a test-negative design and a matched case-control design (Methods).

## Methods

### Data collection and study population

Anonymized Electronic Medical Records (EMRs) were retrieved from MHS’ centralized computerized database. MHS is a 2.5-million-member, state-mandated, non-for-profit, second largest health fund in Israel, which covers 26% of the population and provides a representative sample of the Israeli population. The study population consisted of MHS members, aged 40 and above, who received either two or three doses of the BNT162b2 vaccine. Analyses focused on the period from January 2021 (when the second dose was first administered among eligible individuals) to August 21, 2021, as the booster dose was widely administered from August 1 onwards. Participants were excluded from the study if they disengaged from MHS for any reason prior to March 2020 or if they tested positive for SARS-CoV-2 by a polymerase chain reaction (PCR) test before the start of the follow-up period.

Individual-level data for the study population included age (in 10-year age categories), biological sex, socioeconomic status (SES) index, and a coded geographical statistical area (GSA; the smallest geostatistical unit assigned by Israel’s National Bureau of Statistics, which correspond to neighborhoods). The SES index was measured on a scale from 1 (lowest) to 10 based on several parameters including household income, educational qualifications, household crowding, and car ownership. Data collected also encompassed the last documented body mass index (BMI) (where obesity was defined as BMI ≥ 30) and information on chronic diseases from MHS’ automated registries, including cardiovascular diseases,^14^ hypertension,^15^ diabetes,^16^ chronic kidney disease (CKD),^17^ chronic obstructive pulmonary disease (COPD), and immunocompromised conditions, as well as data on cancer from the National Cancer Registry.^18^ COVID-19-related information included dates of the first, second, and third dose of the vaccine (if received) and results of any PCR tests for SARS-CoV-2, including tests performed outside of MHS.

### Statistical analysis

#### Main analysis: test-negative design

For the main analysis, we used a test-negative design,^19,20^ where ‘cases’ were defined as those who had a positive PCR test for SARS-CoV-2 and ‘controls’ were those testing negative. Individuals could contribute multiple negative tests to the analysis but were excluded once they tested positive. If multiple tests were conducted within a 5-day period, the day of test was considered to be the first day of that test series. In a sensitivity analysis we included all tests regardless of serial testing, and the results were unchanged. The analysis sought to estimate the reduction in the odds of a positive test at different time intervals following receipt of the booster (third) dose (0-6 days, 7-13 days, 14-20 days) compared to two-dose only vaccinees. The 7-day and 14-day cutoffs for the third dose were chosen based on second-dose research.^3,21^ Covariates included the 10-year age category, biological sex, time since receipt of the 2^nd^ dose (time category), and comorbidities (diabetes, immunosuppression status, BMI ≥ 30, CVD). An additional covariate consisted of the number of positive tests perfomed on that day througout the population (log-transformed), attempting to adjust for variability of levels of exposure at different time points. In sensitivity analyses, we evaluated two alternative ways to adjust for time-varying infection risk, by using a covariate consisting of the number of positive tests per day stratified by socioeconomic level, or by using a series of dummy variables representing calendar week. These alternatives yielded estimates of vaccine effectiveness that were similar to the results presented in the main analysis. SES category, COPD, hypertension, and CKD were also tested in the model but were not significant and not included in the final model; their inclusion/exclusion did not influence the estimates of the effect of the booster dose.

A logistic regression model was fit to the data. The marginal benefit of the third dose (compared to those receiving two doses) was calculated as 100%*(1-(Odds Ratio)) for each of the booster dose time-categories.

#### Secondary analysis: matched case-control design

As a complementary analysis, we used a matched case-control design.^22,23^ Cases were defined as individuals with a positive PCR test occurring after August 1, 2021, among those 40 years of age or older who did not have a previous positive test recorded and who received at least two doses of the vaccine. Up to 20 controls per case were drawn from the entire population (64% of the population were matched to 20 controls, 81% of the population had at least 10 controls, and 90% of the population had at least 5 controls). Eligible controls were individuals who had not tested positive (i.e., with either no positive PCR tests or negative tests) before the date when the case was tested and who had received at least two doses of the vaccine. Controls were matched by 10-year age category, residential socioeconomic status, biological sex, and the month when the second dose was administered. Both conditional and unconditional logistic regression models were fit to the data,^24^ and the covariates were the same as described for the test-negative analysis, except socioeconomic status and biological sex, which were not significant and were dropped from the analysis.

Analyses were performed using R version 4.0.5.

### Ethics declaration

This study was approved by the MHS (Maccabi Healthcare Services) Institutional Review Board. Due to the retrospective design of the study, informed consent was waived by the IRB and all identifying details of the participants were removed before computational analysis.

### Data availability statement

According to the Israel Ministry of Health regulations, individual-level data cannot be shared openly. Specific requests for remote access to de-identified community-level data should be referred to KSM, Maccabi Healthcare Services Research and Innovation Center.

### Code availability statement

Specific requests for remote access to the code used for data analysis should be referred to KSM, Maccabi Healthcare Services Research and Innovation Center.

## Results

From August 1, when the booster dose became widely available, through August 21, 2021, 182,076 PCR tests were performed among 153,753 MHS members over the age of 40 who did not have a previous documented infection. Baseline characteristics of the participants are given in Supplementary Table 1 for tests performed during August. During the period when the booster was available, 8,285 positive tests and 149,379 total tests (5.5%) were in the two-dose group and 1,188 positive tests and 32,697 total tests (3.6%) in the three-dose group. The percent positive was highest in those who had not not received a booster and in those who had received a booster in the previous 7 days, and was lowest in individuals who had received the booster more than two weeks prior (Table 1).

**Table 1.**
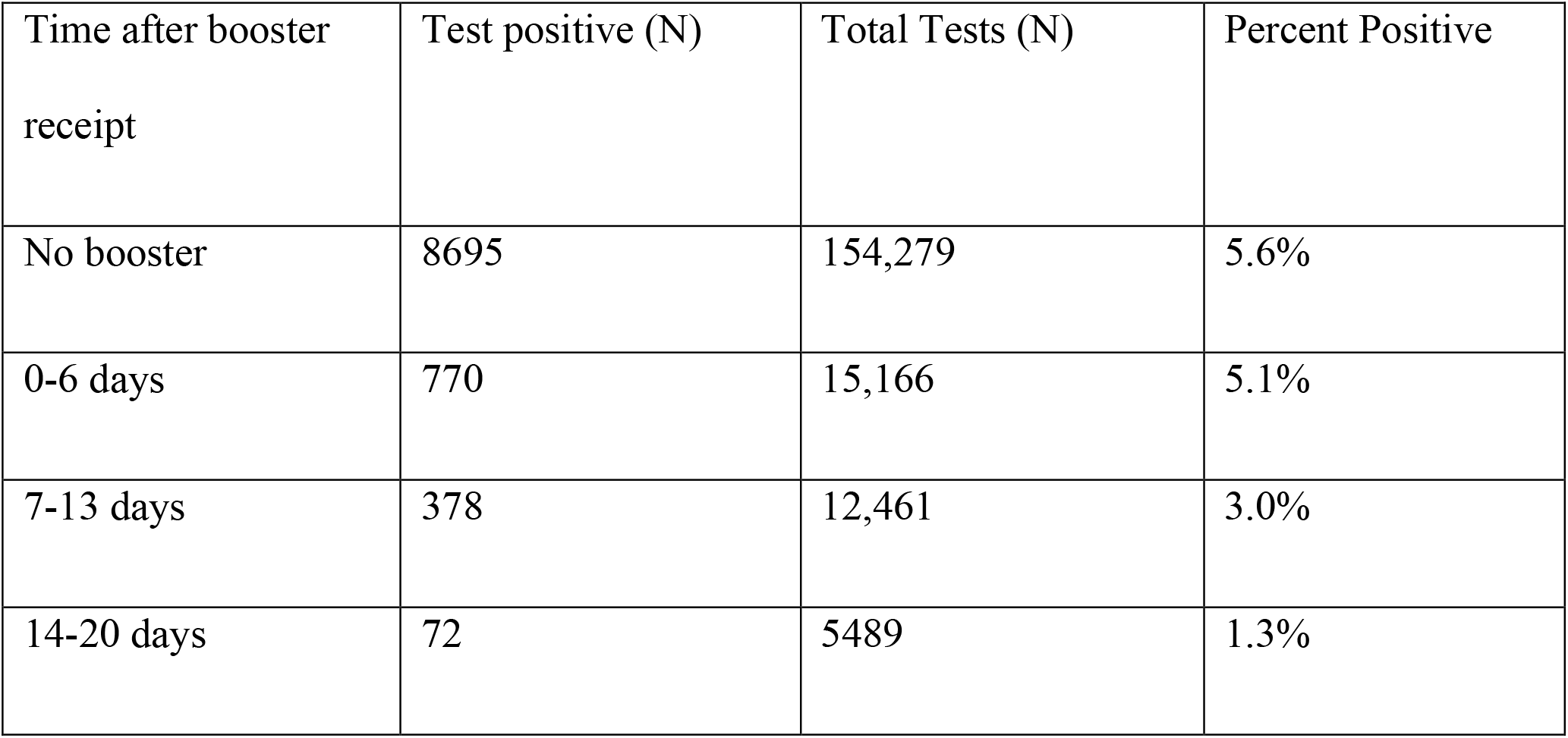
Testing results among those with those with at least two doses of the vaccine at different time points, August 1- August 21, 2021

The marginal effectiveness of the third dose compared to two doses increased over time following receipt of the booster, with no marginal effectiveness in the first 7 days (3%; 95% CI: -5%, 10%), moderate marginal effectiveness in days 7-13 (48%; 95% CI: 42%, 54%), and high marginal effectiveness in days 14-20 (79%; 95% CI: 72%, 84%) (Supplementary Tables 2 and 3, Figure 1).

**Figure 1.**
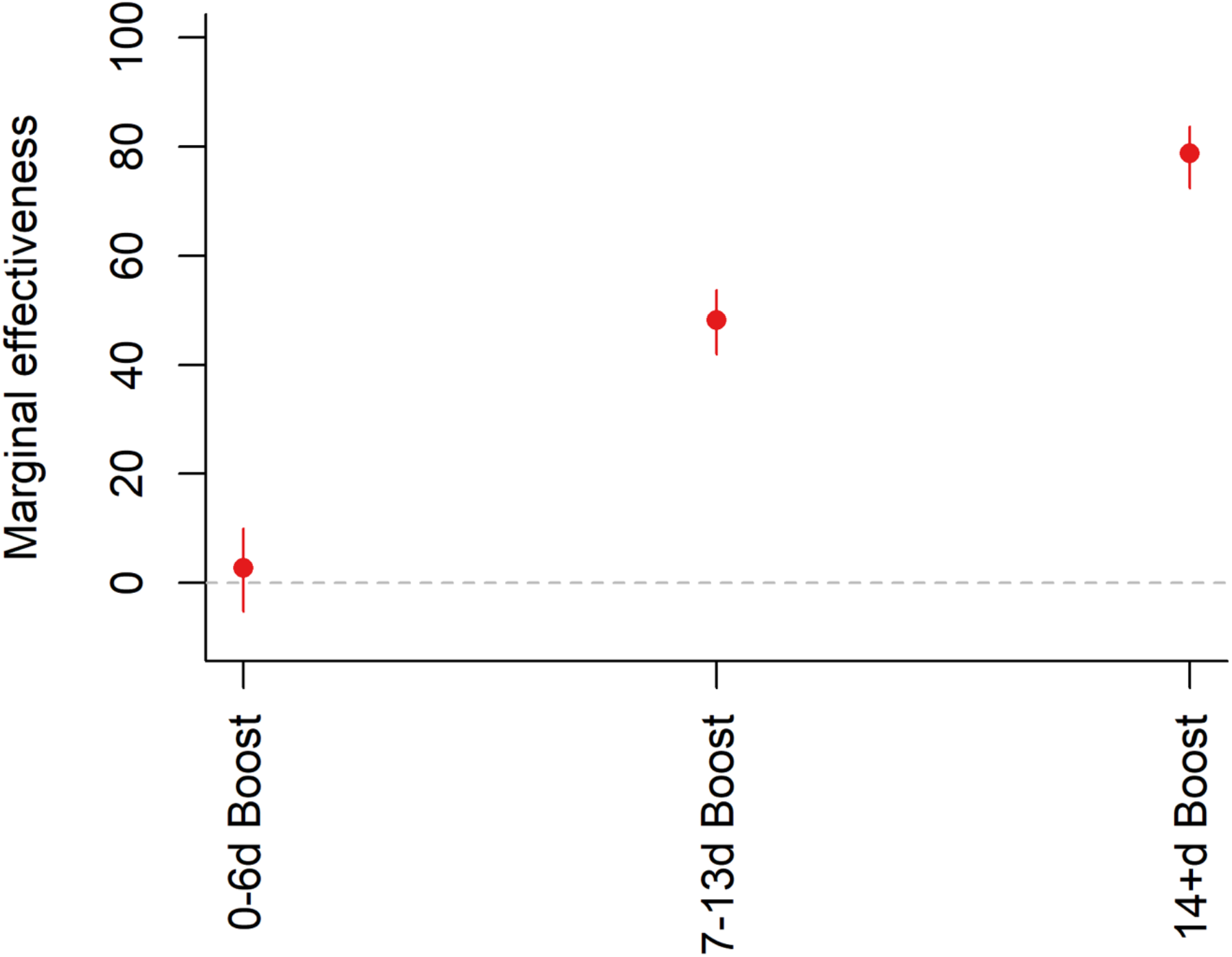
Reduction in the odds of testing positive to SARS-CoV-2 in third-dose vaccinees (marginal effectiveness of the booster).

Similar to the results from the test-negative analysis, the matched case-control analsysis estimated the marginal effectiveness of the booster dose compared to the second dose increased from 39% (95% CI: 34%, 44%) 0-6 days after the booster to 53% (95% CI: 48%, 58%) 7-13 days after the booster and 70% (95% CI: 62%, 76%) 14-20 days after the booster (Supplementary Tables 2 and 4). A conditional logistic regression model gave qualitatively similar results (Supplementary Table 2).. Baseline characteristics of the participants are given in Supplementary Table 5.

## Discussion

In this study, we found that a third dose of the BioNTech/Pfizer mRNA BNT162b2 vaccine provided additional protection against detected infection. Across the test-negative and matched case-control analyses, we estimated a 48-68% reduction in the odds of testing positive for SARS-CoV-2 after 7-13 days and 70-84% 14-20 days after the booster compared to two doses.

Our study has the inherent limitation of being short-term and preliminary, and we only evaluate marginal effectiveness against infection rather than severe disease. The total benefit of the vaccine program will depend both on the long-term effectiveness of the first two doses of the vaccine, accounting for waning, and the marginal improvements conferred by receipt of the booster. These analyses focused on infections, which accrue quickly and in high numbers. As more time elapses, it will be possible to evaluate the effects on more rare but severe outcomes, such as hospitalization and death. It will also be important to monitor waning of effectiveness following receipt of the booster dose. There are methodological limitations to these analyses as well. As in every observational study, there is a potential bias relating to healthcare seeking behavior in terms of PCR test frequency in the examined groups.

This could account for the discrepancy of the results in days 0-6 between the matched case-control analysis and the test-negative analysis. The test-negative design attempts to mitigate that potential bias but it cannot eliminate it. As those eligible for the third dose are early vaccinees, and given that chronically-ill patients were prioritized for vaccination, uncontrolled confounding by indication is possible. However, the matching and adjustment perfomed renders residual confounding by unmeasured factors less likely. We use odds ratios as an approximation of the relative risk and marginal effectiveness of the third dose, which is a reasonable approximation given that outcome is relatively rare. Another limitation stems from the fact that the Delta variant is currently the dominant strain in Israel, thus the risk reduction against other strains cannot be inferred.

In conclusion, other studies have demonstrated that protection of vaccines against SARS-CoV-2 wanes over time. Our analyses show that the waning of vaccine-induced protection against infection, seems to be counteracted in the short term by the third dose. Further monitoring of data from this population is needed to determine the duration of immunity following the booster.

## Supplementary Tables and Figures

**Supplementary Table 1.** Demographic characteristics associated with tests among individuals with at least 2 doses of vaccine who were tested August 1-21, 2021

| Characteristics | Two-dose only vaccinees<br>(n=149,379) | Three-dose vaccinees<br>(n=32,697) |
| --- | --- | --- |
| **Age years, mean (SD)** | 53.6 (10.9) | 68.2 (10.7) |
| **Age group – no. (%)** |  |  |
| 40 to 49 yr | 65,517 (43.9%) | 712 (2.2%) |
| 50 to 59 yr | 50,072 (33.5%) | 4865 (14.9%) |
| 60 to 69 yr | 19,029 (12.7%) | 13,603 (41.6%) |
| 70-79 yr | 9,441 (6.3%) | 8320 (25.4%) |
| ≥80 yr | 5320 (3.6%) | 5197 (15.9%) |
| **Sex – no. (%)** |  |  |
| Male | 65,474 (43.8%) | 15,198 (46.5%) |
| Female | 83,905 (56.2%) | 17,499 (53.5%) |
| **SES, mean (SD)** | 6.9 (1.8) | 6.9 (1.9) |
| **Comorbidities – no. (%)** |  |  |
| CVD | 2341 (1.6%) | 1647 (5.0%) |
| Obesity (BMI ≥30) | 33,482 (22.4%) | 9259 (28.3%) |
| **Note:** this table is related to the number of tests; individuals can be counted more than once for these calculations |  |  |

SD – Standard Deviation; SES – Socioeconomic status on a scale from 1 (lowest) to 10; CVD – Cardiovascular Diseases; BMI – Body Mass Index.

**Supplementary Table 2.** Marginal effectiveness of 2 doses versus 3 doses

| Time after booster | Test-negative analysis | Matched case-control<br>(unconditional) | Matched case-control<br>(conditional) |
| --- | --- | --- | --- |
| 0-6 days | 3% (-5%, 10%) | 39% (34%, 44%) | 48% (43%, 52%) |
| 7-13 days | 48% (42%, 54%) | 53% (48%, 58%) | 68% (64%, 72%) |
| 14-20 days | 79% (72%, 84%) | 70% (62%, 76%) | 84% (79%, 88%) |

**Supplementary Table 3.** OR for SARS-CoV-2 infection, demographics and comorbidities, test-negative design, two dose vaccinees vs. third-dose vaccinees

| Variable | Category | OR (95% CI) |
| --- | --- | --- |
| **Vaccination status** |  |  |
|  | Two dose vaccinees | Ref |
|  | Three-dose vaccinees, days 0-6 | 0.97 (0.90, 1.05) |
|  | Three-dose vaccinees, days 7-13 | 0.52 (0.46, 0.58) |
|  | Three-dose vaccinees, days 14+ | 0.21 (0.16, 0.28) |
| **Age group, yr.** |  |  |
|  | 40-49 | Ref |
|  | 50-59 | 0.91 (0.87, 0.93) |
|  | 60-69 | 0.88 (0.84, 0.93) |
|  | 70-79 | 0.90 (0.85, 0.96) |
|  | ≥80 | 0.69 (0.64, 0.75) |
| **Sex** |  |  |
|  | Male | Ref |
|  | Female | 0.8 (0.77, 0.83) |
| **Comorbidities** |  |  |
|  | Obesity (BMI≥30) | 1.19 (1.15, 1.24) |
|  | Diabetes mellitus | 1.23 (1.17, 1.29) |
|  | Immunosuppression | 1.11 (1.02, 1.21) |
|  | Cardiovascular diseases | 1.16 (1.04, 1.28) |

OR – Odds Ratio

**Supplementary Table 4.** OR for SARS-CoV-2 infection, demographics and comorbidities, matched control desgin, two dose vaccinees vs. third-dose vaccinees, unconditional logistic regression

| Variable | Category | OR |
| --- | --- | --- |
| **Vaccination status** |  |  |
|  | Two dose vaccinees | Ref |
|  | Three-dose vaccinees, days 0-6 | 0.61 (0.56, 0.66) |
|  | Three-dose vaccinees, days 7-13 | 0.47 (0.42, 0.52) |
|  | Three-dose vaccinees, days 14+ | 0.30 (0.24, 0.38) |
| **Age group, yr.** |  |  |
|  | 40-49 | Ref |
|  | 50-59 | 1.02 (0.98, 1.06) |
|  | 60-69 | 1.17 (1.11, 1.23) |
|  | 70-79 | 1.21 (1.14, 1.28) |
|  | ≥80 | 1.48 (1.37, 1.61) |
| **Sex** |  |  |
|  | Male | Ref |
|  | Female | 0.97 (0.94, 1.00) |
| **Comorbidities** |  |  |
|  | Obesity (BMI≥30) | 1.12 (1.08, 1.16) |
|  | Diabetes mellitus | 1.1 (1.05, 1.16) |
|  | Immunosuppression | 1.35 (1.24, 1.46) |
|  | Cardiovascular diseases | 1.23 (1.11, 1.37) |

OR – Odds Ratio

**Supplementary Table 5.** Characteristics of participants, matched case control analysis.

| Characteristics | Two-dose only vaccinees<br>(n=238,018) | Three-dose vaccinees<br>(n=30,295) |
| --- | --- | --- |
| **Age years, mean (SD)** | 54.2 (11.0) | 66.2 (9.46) |
| **Age group – no. (%)** |  |  |
| 40 to 49 yr | 101,900 (42.8%) | 559 (1.2%) |
| 50 to 59 yr | 75,337 (31.7%) | 6,272 (20.7%) |
| 60 to 69 yr | 34,298 (14.4%) | 12,991 (42.9%) |
| 70-79 yr | 18,855 (7.9%) | 7,946 (26.2%) |
| ≥80 yr | 7,628 (3.2%) | 2,527 (8.3%) |
| **Sex – no. (%)** |  |  |
| Male | 115,276 (48.4%) | 17,121 (56.5%) |
| Female | 122,742 (51.6%) | 13,174 (43.5%) |
| **SES, mean (SD)** | 6.8 (1.8) | 6.7 (1.8) |
| **Comorbidities – no. (%)** |  |  |
| CVD | 4170 (1.8%) | 1302 (4.3%) |
| Obesity (BMI ≥30) | 56,397 (23.7%) | 8731 (28.8%) |

SD – Standard Deviation; SES – Socioeconomic status on a scale from 1 (lowest) to 10; CVD – Cardiovascular Diseases; BMI – Body Mass Index.

## Notes

### Competing Interest Statement

Conflicts of interest:
DMW has received consulting fees from Pfizer, Merck, Affinivax, and Matrivax for work unrelated to this paper and is Principal Investigator on grants from Pfizer and Merck to Yale University for work unrelated to this manuscript. VEP has received reimbursement from Merck and Pfizer for travel to Scientific Input Engagements unrelated to the topic of this manuscript and is a member of the WHO Immunization and Vaccine-related Implementation Research Advisory Committee (IVIR-AC). JW discloses consulting fees from Revelar Inc.

### Funding Statement

Research reported in this publication was supported by NIAID of the National Institutes of Health under award number R01AI137093. The content is solely the responsibility of the authors and does not necessarily represent the official views of the National Institutes of Health.

## References

1. Polack, F. P. et al. Safety and Efficacy of the BNT162b2 mRNA Covid-19 Vaccine. N. Engl. J. Med. 383, 2603–2615 (2020).

2. Baden, L. R. et al. Efficacy and Safety of the mRNA-1273 SARS-CoV-2 Vaccine. https://doi.org/10.1056/NEJMoa2035389 384, 403–416 (2020).

3. Dagan, N. et al. BNT162b2 mRNA Covid-19 Vaccine in a Nationwide Mass Vaccination Setting. N. Engl. J. Med. 384, (2021).

4. Chodick, G. et al. The Effectiveness of the Two-Dose BNT162b2 Vaccine: Analysis of Real-World Data. Clin. Infect. Dis. (2021) doi:10.1093/CID/CIAB438.

5. Seow, J. et al. Longitudinal observation and decline of neutralizing antibody responses in the three months following SARS-CoV-2 infection in humans. Nat. Microbiol. 5, 1598–1607 (2020).

6. Ruopp, M. D., Strymish, J., Dryjowicz-Burek, J., Creedon, K. & Gupta, K. Durability of SARS-CoV-2 IgG Antibody Among Residents in a Long-Term Care Community. J. Am. Med. Dir. Assoc. 22, 510–511 (2021).

7. Shrotri, M. et al. Spike-antibody waning after second dose of BNT162b2 or ChAdOx1. Lancet 0, (2021).

8. Mizrahi, B. et al. Correlation of SARS-CoV-2 Breakthrough Infections to Time-from-vaccine; Preliminary Study. medRxiv 2021.07.29.21261317 (2021) doi:10.1101/2021.07.29.21261317.

9. COVID-19 in Israel dashboard. (2021).

10. Health Ministry expands COVID booster shot drive to ages 30 and up | The Times of Israel. https://www.timesofisrael.com/health-ministry-expands-covid-booster-shots-to-ages-30-and-up/.

11. Kamar, N. et al. Three Doses of an mRNA Covid-19 Vaccine in Solid-Organ Transplant Recipients. https://doi.org/10.1056/NEJMc2108861 (2021) doi:10.1056/NEJMC2108861.

12. Massa, F. et al. Safety and Cross-Variant Immunogenicity of a Three-Dose COVID-19 mRNA Vaccine Regimen in Kidney Transplant Recipients. SSRN Electron. J. (2021) doi:10.2139/SSRN.3890865.

13. Pfizer and BioNTech Provide Update on Booster Program in Light of the Delta-Variant. https://investors.biontech.de/news-releases/news-release-details/pfizer-and-biontech-provide-update-booster-program-light-delta.

14. Shalev, V. et al. The use of an automated patient registry to manage and monitor cardiovascular conditions and related outcomes in a large health organization. Int. J. Cardiol. 152, 345–349 (2011).

15. D, W., G, C., V, S., C, G. & E, G. Prevalence and factors associated with resistant hypertension in a large health maintenance organization in Israel. Hypertens. (Dallas, Tex. 1979) 64, 501–507 (2014).

16. Chodick, G., Heymann, A. D., Shalev, V. & Kookia, E. The epidemiology of diabetes in a large Israeli HMO. Eur. J. Epidemiol. 18, 1143–1146 (2003).

17. Coresh, J. et al. Decline in estimated glomerular filtration rate and subsequent risk of end-stage renal disease and mortality. JAMA - J. Am. Med. Assoc. 311, 2518–2531 (2014).

18. Israel Center for Disease Control. Jerusalem, I. Data from: Israel national cancer registry.

19. Lewnard, J. A. et al. Theoretical Framework for Retrospective Studies of the Effectiveness of SARS-CoV-2 Vaccines. Epidemiology 32, 508 (2021).

20. Vandenbroucke, J. P. & Pearce, N. Test-negative designs: Differences and commonalities with other case-control studies with ‘other patient’ controls. Epidemiology 30, 838–844 (2019).

21. Chodick, G. et al. The Effectiveness of the Two-Dose BNT162b2 Vaccine: Analysis of Real-World Data. Clin. Infect. Dis. (2021) doi:10.1093/CID/CIAB438.

22. V, E., RW, P., M, A. & L, P. Comparison of nested case-control and survival analysis methodologies for analysis of time-dependent exposure. BMC Med. Res. Methodol. 5, (2005).

23. Zakeri, R. et al. A case-control and cohort study to determine the relationship between ethnic background and severe COVID-19. EClinicalMedicine 28, (2020).

24. Kuo, C.-L., Duan, Y. & Grady, J. Unconditional or Conditional Logistic Regression Model for Age-Matched Case–Control Data? Front. Public Heal. 6, 2 (2018).

